# What can the ideal gas say about global pandemics? Reinterpreting the basic reproduction number

**DOI:** 10.1101/2020.07.09.20150128

**Authors:** Bradley M. Dickson

**Affiliations:** Center for Epigenetics, Van Andel Research Institute, Grand Rapids Michigan 49503, USA

## Abstract

Through analysis of the ideal gas, we construct a random walk that on average matches the standard susceptible-infective-removed (SIR) model. We show that the most widely referenced parameter, the “basic reproduction number” (ℛ_0_), is fundamentally connected to the relative odds of increasing or decreasing the infectives population. As a consequence, for ℛ_0_ > 1 the probability that no outbreak occurs is 1*/*ℛ_0_. In stark contrast to a deterministic SIR, when ℛ_0_ = 1.5 the random walk has a 67% chance of avoiding outbreak. Thus, an alternative, probabilistic, interpretation of ℛ_0_ arises, which provides a novel estimate of the critical population density *γ/r* without fitting SIR models. We demonstrate that SARS-CoV2 in the United States is consistent with our model and attempt an estiamte of *γ/r*. In doing so, we uncover a significant source of bias in public data reporting. Data are aggregated on political boundaries, which bear no concern for dispersion of population density. We show that this introduces bias in fits and parameter estimates, a concern for understanding fundamental virus parameters and for policy making. Anonymized data at the resolution required for contact tracing would afford access to *γ/r* without fitting. The random walk SIR developed here highlights the intuition that any epidemic is stochastic and recovers all the key parameter values noted by Kermack and McKendrick in 1927.

## INTRODUCTION

The past few months have been dominated by the global pandemic caused by SARS-CoV2. The susceptible-infective-removed (SIR) model[1] has become popularized and is frequently seen in the media. Reports are emerging that aim to control misinterpretation of the model and motivate further developments of models, even alternative models, for infectious disease.[2–6]

Estimation of critical values, those defining the likelihood of an outbreak, are reguarded as difficult to estimate for various reasons.[7] As we build increasingly complex models to better fit empirical data and estimate crititcal parameters, one has to also manage the extent to which the models correctly account for ‘the physics’ underlying the empirical data. With this in mind, our current goal is to enable evaluation of critical parameters directly from empirical data with as few assumptions as possible. In the end, our key assumption is that we have access to data that can indicate the population density at which illness causing exposures occurred. Recent projects are emerging with the hopes of making this data a reality.[8] At the moment however, we do not have access to such data.

Toward our aim, this article introduces a version of the SIR model from the perspective of statistical mechanics. We show that the ideal gas system, a common benchmark in statistical physics, can give rise to a random walk that on average recovers all the results of the standard SIR model. The random walk leads to new interpretations of ℛ_0_, the “base reproductive number.” Importantly, we find ℛ_0_ as the ratio of instantaneous probabilities for increasing or decreasing the number of infectives and therefore it controls the odds of outbreak for fixed population density. As a mild departure from the deterministic statement that ‘outbreak will occur if ℛ_0_ > 1’ we find that an outbreak will occur with probability 1 − 1*/*ℛ_0_. We demonstrate that SARS-CoV2 data in the United States likely conforms to this interpretation but our ability to accurately evaluate threshold values is limited by the fact that publicly available data is aggregated on taxation and political boundaries, disregarding population dispersion. We note that the ideal gas has appeared in previous literature for epidemic modelling, though we take a different route of analysis here.[9]

In what follows we will introduce the standard SIR model and restate its key observations. We then define a random walk that is on average consistent with the standard model. We present the average number of initial secondary infections, the average time-to-peak infection, and a novel observation on ℛ_0_ regarding the odds of outbreak. Finally, we consider applying the results to the United States SRAS-CoV2 data.

## THEORY AND RESULTS

### Standard SIR model

Following the language of Kermack and McKendrick[1] we define the susceptible (*S*), infective (*I*), and removed (*R*) populations. The population *S* are the members of the population that are not immune to the disease in question. Like Kermack and McKendrick we assume that is the whole population. The infective population *I* are those currently infected and actively spreading the disease. The removed *R* population are those who have recovered or succumbed to the illness, or have simply decided to stay home while sick so as to mitigate spread. In what follows all of these populations are in fact population densities. It is clear on inspection of equation 4 in reference 1 that Kermack and McKendrick also formulated their model in terms of population density. In a fixed area, we have the conservation law *N* = *S* + *R* + *I* = *S*_0_ + *I*_0_ where the total population density *N* is fixed at all time. *S*_0_ and *I*_0_ are the initial values for suceptible and infective. The initial removed population is always set to naught.

Kermack and McKendrick noted that there is a critical density *N* ^*^. Densities below *N* ^*^ do not experience full outbreak if exposed to one (*I*_0_ = 1) initial infection. Densities above *N* ^*^ will experience an epidemic, the severity of which increases with the density until the population is fully saturated.

The SIR model for the dynamics of an outbreak, under the assumption of constant rates of infectivity *r* and removal *γ*, is

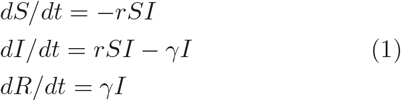

In the present, the initial values for this system of equations are always understood to be *S*_0_ = *N* − 1, *I*_0_ = 1, and *R*_0_ = 0. The critical density found by Kermack and McKendrick was *N* ^*^ = *γ/r*. The units of *r* are then [*dt*]^−1^[*N*]^−1^, which is reciprocal unit time by reciprocal unit density. In a discrete system *SI* is the number of pairs between members of the *S* and *I* populations, so *r* can be seen as setting the fraction of those pairs that will transition from status *S* to status *I* in a unit time. The *S* and *I* are continuous in the standard SIR model, however. The parameter *γ* has units of [*dt*]^−1^ and sets the rate of leaving status *I* for status *R*. The widely known “base reproduction number” (or ratio), ℛ_0_, is ℛ_0_ = *rS*_0_*/γ* and is observed by considering *dI/dt*. Our definition is consistent with Daley and Gani, see problem 1.2 on page 17 for example.[10] We note that interpretation of ℛ_0_ is a source of some debate[11, 12]. If *dI/dt <* 0, *rS/γ <* 1 and the infectives do not expand in number. If *dI/dt* > 0, then *rS/γ* > 1 and the infectives do expand in number. The critical density is *N* ^*^ = *γ/r* which corresponds to ℛ_0_ = 1.

The number of infectives generated by the first lone infective, in the first unit time, is *rS*_0_. On average it takes 1*/γ* time units to remove this initial infective. Thus, ℛ_0_ = *rS*_0_*/γ* is the average number of infectives created by the initial infective assuming that ℛ_0_ is a negligible fraction of *S*_0_ so that there is a constant rate of infection during the 1*/γ* time interval.

The above model is formulated on continuous variables *S, R*, and *I* and with a more or less macroscopic notion of the individuals in the population. At any instant of time the rate of change of *S* is −*rSI* which need not be an integer nor be greater than unity depending on the choice of unit time and *r*. What we consider next is how to define the SIR model wherein each individual of the population is explicit and the change −*rSI* is limited to integer values. Thus, in a unit time the number of susceptible *S*, infective *I* and removed *R* change by integers as they would in the “real world.” Ultimately, treating the population as individuals would allow a number of advances toward more realistic particle-based modelling. Contact tracing, sub-populations with poor mixing, variations in dwell times, etc., could be studied in a full particle-based model.

### Ideal gas

Consider an ideal gas model of a particle system in the canonical ensemble with fixed number of particles, fixed area containing the particles, and fixed temperature *T*. In the ideal gas, the particles spread randomly with uniform probability density over the area they occupy and they exert no force on one another. Each particle carries with it a small circular boundary defining the radius of contact. If another particle is found within this boundary, the two particles are considered to be in contact. We use *A*_*i*_ to indicate the *i*-th particle’s area of contact and we write *A* for the total area. All the radii of contact are equal.

One can simulate all the particles in the system, just as is done in molecular dynamics simulations. The simulation would solve two stochastic differential equations, two Langevin equations, for each particle to obtain the change in particle position as a function of time. Disease transmission can be modelled in this system by including an attribute for each particle. The attribute is just a label, *S, I*, or *R* which indicates whether the particle is susceptible, infective, or removed. If one particle with label *S* is found in contact with a particle labeled *I*, we can model probabilistic disease transmission with a “coin toss.” We assign the probability of transmission to be *p*_*T*_ and let the “coin” come up heads with *p*_*T*_ probability. If one *S* particle is in contact with an *I* particle, the coin toss is performed and the *S* attribute transitions to *I* if the coin is heads. The label change does not impact the dynamics of the particles at all, it is only a label.

Rather than do the full particle simulations, we developed expressions for the dynamics of this system. This reduces simulation time, by avoiding the simulation, and ultimately allows us to treat millions of particles extremely rapidly. The particle simulations, we know, will sample the Boltzmann distribution *ρ* ∝ exp[−*H/k*_*B*_*T*] with *H* the system Hamiltonian and *k*_*B*_ the Boltzmann constant. So we may approach this problem from the stand point of statistical mechanics in the canonical ensemble.[13]

Let there be one infected particle, say the *l*-th particle, and set the number of infected particles to *I* = 1. For the ideal gas, the probability that a number *x* of the *S* particles are in contact with the one infected particle, can be evaluated with the following configuration integral[14]

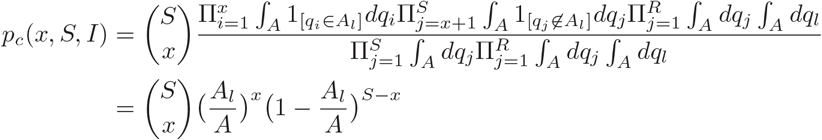

The indicator function 1_[·]_ is unity if its argument is satisfied and zero otherwise. The momenta integrate away in this case and we let *q*_*i*_ denote the 2-dimensional position of the *i*-th particle. Integrals are understood to be over the fixed area occupied by the particles. In the ideal gas, increasing infectives in turn increases the area that is potentially infectious. If there are 3 infectives, the probability of contacting an infective goes up three fold. The probability of contacting one infective is *A*_*l*_*/A* and so the odds of contacting any of the three infectives is 3*A*_*l*_*/A*. When the contact radius is measured in square feet and the containing area in square miles, as is the case here, the particle-particle contact area *A*_*i*_ is sufficiently small compared to the total area *A* so that *IA*_*i*_*/A* ≤ 1 even if *I* = *N*. If the probability of disease transmission upon contact is *p*_*T*_ *<* 1, then the joint probability of contacting a single infective and contracting illness is *r* = *p*_*T*_ *A*_*l*_*/A. r* is the odds of contracting illness *per infective*. Thus, the parameters of the standard SIR model are mixing what we consider fundamental measurements of the illness (*p*_*T*_) and attributes of the population (*A*_*l*_*/A*). Sampling *r* from a distribution then can be viewed as a coarse-grained treatment of behavioral differences in individuals of the population. The total probability of contracting *x* new illnesses is then

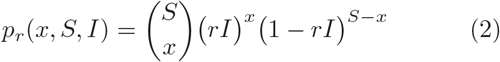

This is just the binomial distribution, as expected for the ideal gas.[14] The binomial distribution can give rise to a Poisson distribution when *S* is very large compared to *x* and *λ* = *rIS*. Here the Poisson parameter will be a function of *I*, as intuitively captured by the ideal gas. The expected number of new infections for this system with *S* susceptible and *I* infectives is

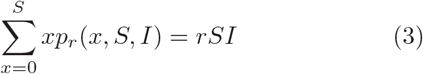

This result shows that on average, the ideal gas system with *S* susceptible and *I* infective will generate the same number of new infectives *rSI* as the standard SIR model of equation (1). In Reference 1 equation 29, *κ* is *r* here. We have explicitly connected the parameter *r* to the probability of contact in the particle system *A*_*l*_*/A. r* contains information about the total area confining the population as well as information about the contact distance. In what follows our contact area is set as a circle of a seven foot radius and the total area is one square mile, *A*_*i*_*/A* = 6 × 10^−6^. We also arbitrarily take *p*_*T*_ = 0.1 to assign a 10% chance of contracting illness on infective contact. The above results place physical interpretations and limitations on *r*.

A note about “time.” The ideal gas is at equilibrium and the only things changing are the *S, I, R* labels but these labels do not alter the dynamics (or distribution) of the gas particles. We can then invent a “unit time” by stating that this unit amount of time has passed each time we evaluate equation (2). If we were to perform molecular dynamics simulations of the ideal gas, evaluating equation (2) amounts to observing the system configuration and reassigning the *S, I, R* labels according to our contact and transmission criteria. The next observation and label exchanges can take place on any timescale longer than the dynamical correlation time. In our case we will evaluate equation (2) every unit time *τ* = 1 rather than do explicit simulations of the ideal gas. This time unit could be calibrated to real data recorded by matching timescales. The time passing before the first infective is removed can be used to tune *γ*. Obviously real world situations are more complex as we expect *γ* to change as awareness of deadly disease heightens. Nevertheless, we henceforth absorb *τ* in *r* which becomes the odds of contracting illness *per infective per unit time*. Consistent with the standard SIR we assume interactions last for some average length of time.

For the *I* infective particles we may also define the probability that any one (or more) of them will transition to the removed set

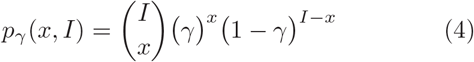

Again consistent with the SIR model, the instantaneous number of particles transitioning to the removed set in a unit time is *γI*. Like *r* above, we let *γ* absorb the arbitrary unit time to represent the odds of removing an infective per unit time. In terms of the ideal gas, this is treated as a change of label only and does not impact the dynamics of the gas. With respect to configuration integrals, the process of transitioning to removed matches the probability of finding infected particles in an area of size *γA*. Once an infected particle hits this area, it becomes “removed.” So far we have an ideal gas that on average behaves as the SIR model with respect to *r* and *γ*.

### A random walk for S, I, and R

Consider building a random walk of the label/attribute updates based on *p*_*r*_(*x*) and *p*_*γ*_(*x*). We define the random variables

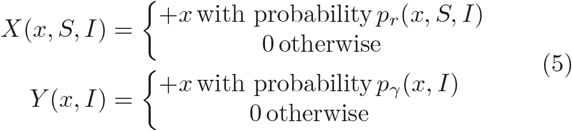

Let 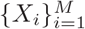 be a sequence of independent observations of *X* for a fixed *x, S*, and *I*. Then 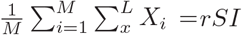 for large *M* and *L*. For the random variable *Y* we have a similar result. Thus, the random walk is 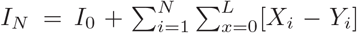 is comprised of increments 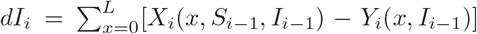 that on average increase infectives by *rSI* while decreasing infectives on average by −*γI* at each unit time. Yet, the susceptible, infective, and removed populations change by integer counts.

The *M* -th step of the random walk is

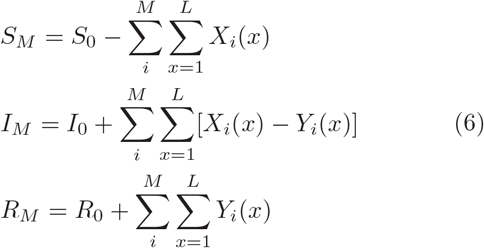

where *τ* time passes at each update. Notice that if we average the change in infectives at the first step

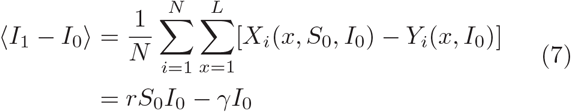

If ⟨*I*_1_ − *I*_0_⟩ > 0, then *rS*_0_*/γ* > 1. If ⟨*I*_1_ − *I*_0_⟩ *<* 0, then *rS*_0_*/γ <* 1. If we define ℛ_0_ = *rS*_0_*/γ*, then we arrive at the expected conclusions: If ℛ_0_ > 1 infectives will increase (outbreak), and if ℛ_0_ *<* 1 infectives will decrease (no outbreak). For the random walk however, this is only true on the average. It would not be impossible to see no outbreak even if R_0_ > 1 and vice versa. We make these statements more concrete in the next subsection.

On average, the random walk generates its first secondary infective in

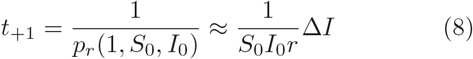

iterations, Δ*I* = 1 new infective. The initial infective, *I*_0_, generates its second infective after another

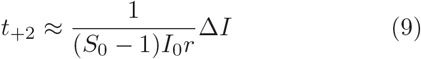

iterations. We can build *t*_+3_ and so on. The number of iterations to, on average, generate *n* infectives is

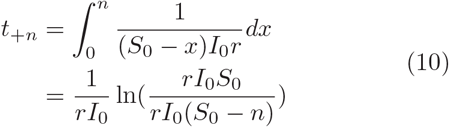

*I*_0_ appears here to make the units clear, but *I*_0_ = 1. We can solve for the number of secondary infectives, *n*_*γ*_, generated by *I*_0_ after 1*/γ* iterations to find

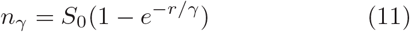

Expanding the exponential to first order shows that *I*_0_ generates *n*_*γ*_ = *rS*_0_*/γ* new infectives. The standard SIR model suggests *I*_0_ will generate ℛ_0_ = *rS*_0_*/γ* infectives before being removed and we have just obtained the same result for the random walk.

### A new interpretation of ℛ_0_

For the random walk of equation (6) we can express the relative odds that infectives, *I*_*M*_, will next increase or decrease by an amount *x* as

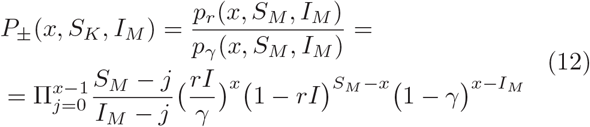

Importantly, when 1 − *rI* ∼ 1 and 1 − *γ* ∼ 1,

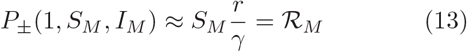

More specifically, this gives us

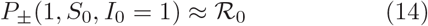

Thus the widely used base reproduction number, ℛ_0_, is equal to the ratio of odds that the next update to equation (6) will increase or decrease by one infective. The inverse function 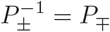 is also a useful quantity. When *P*_±_ > 1, *P*_∓_(1, *S, I*) is the fraction of all realizations of the random walk at *I*_*M*_, *S*_*M*_ that will decrease in the next non-zero update.

The equivalence *P*_±_(1, *S*_0_, *I*_0_ = 1) = ℛ_0_ provides a new interpretation of ℛ_0_. If ℛ_0_ *<* 1 the process *I*_*M*_ is more likely to remove the initial infective in the first non-zero update than it is to generate new infectives. When ℛ_0_ > 1 the process *I*_*M*_ is more likely to generate a new infective in the first non-zero update than it is to remove the initial infective. The generation of a new infective is equally likely as the removal of the initial infective on a non-zero update when ℛ_0_ = 1. On average then, min[1, 1*/*ℛ_0_] is the fraction of realizations of *I*_*M*_ that will terminate without significant increase beyond the initial infective for any ℛ_0_ > 1. The fraction of realizations that end before a time *t*^*^ will be called ℱ(*t < t*^*^). We illustrate this result in Figure 1A, showing that the fraction of realizations ending early without widespread outbreak are given by 1*/*ℛ_0_. Recently extreme value theory has been motivated as a tool for studying epidemics and here we have shown the odds of escaping outbreak follow a “fat-tail” distribution.[5]

**FIG. 1:**
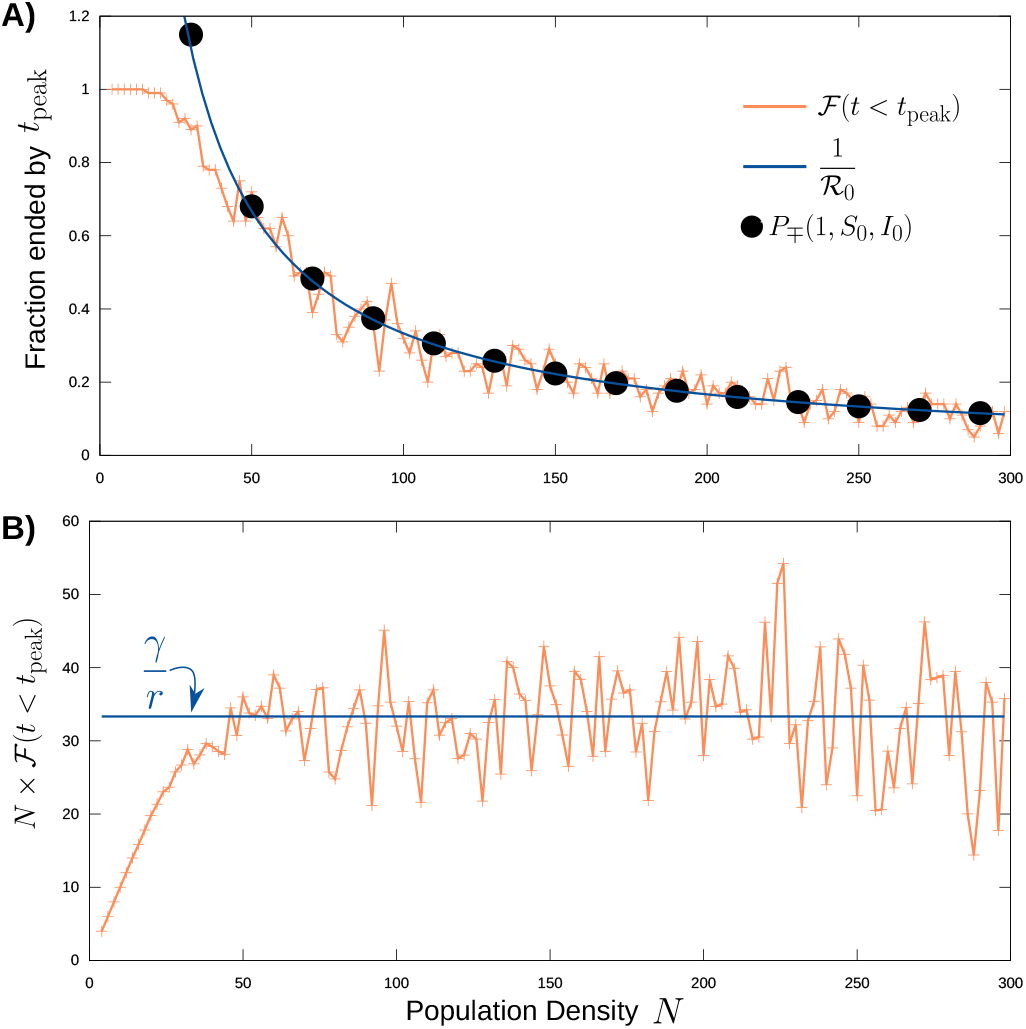
Fraction of early outbreak termination: 100 random walks were generated at different *N* with *r* = 6×10^−7^ and *γ* = 0.00002. (A) The fraction of walks terminated by *t*_peak_, ℱ(*t < t*_peak_) with *t*_peak_ is defined in equation (17), plotted with *P*_±_(1, *S*_0_, *I*_0_) and 1*/*ℛ_0_. (B) Critical density *γ/r* is estimated from ℱ(*t* < *t*_peak_).

The standard SIR model concludes there will be an outbreak when *dI/dt* = *rSI* − *γI* > 0. This condition for growth of the infectives leads directly to the conclusion that ℛ_0_ = *rS*_0_*/γ* > 1 leads to outbreak. The stochastic interpretation of this condition is that *there may be an outbreak* with odds not greater than 1−1*/*ℛ_0_ for ℛ_0_ > 1. When ℛ_0_ = 1.5, the odds of outbreak are not more than 33%. For the standard SIR, though, the conclusion is not probabilistic and thus there must be growth in infectives. The random walk appreciates that while it is 1.5 times more likely to increase infectives it is not impossible nor even all that unlikely to see instead a reduction in infectives. In the initial phase of an epidemic the number of infectives is small and thus at early times it is likely that these infectives will be removed before causing massive spread even if ℛ_0_ > 1. Stochastic effects are known to elevate the threshold of outbreak relative to the deterministic case, and our findings are consistent with this.[7]

### Estimating time to peak infectives

The random walk model presents two timescales. The set of walks that end without outbreak terminate at early times. The walks that increase initial infectives end at late times, but only after consuming enough of the susceptible population that *S*_*M*_ ∼ *γ/r* so that *P*_±_ ∼ 1. When this threshold is met, decreasing infectives and increasing infectives are equally likely (equation (13)) and the momentum for new infectives breaks. We define the critical population 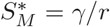 for which *P*_±_ = 1. Once *S*_0_ − *γ/r* susceptibles have been removed from *S*_0_ the epidemic will peak and removal of infectives will become more likely than generation of new infectives. This critical threshold is the same as found in reference 1, 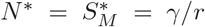, see the text following equation 32 there. Figure 1B demonstrates that where ℱ(*t < t*^*^) can be measured, it affords an estimate of the critical value *γ/r* = *N* × ℱ(*t < t*^*^).

Transitions out of the susceptible state are independent from transitions into the removed state and we have equation (12) for the likelihood of new infectives. From this likelihood we can estimate the average number of iterations required to gain one infective from the initial conditions as

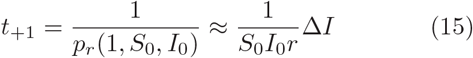

where Δ*I* = 1 is the new infective. The average number of iterations to generate an additional new infective is

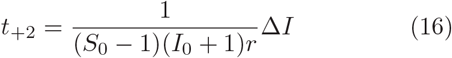

and so on. The epidemic spread breaks momentum when the susceptible population reaches the threshold *γ/r*, as noted above. The cumulative number of infectives, which also counts those infectives that have transitioned to removed, is therefore *S*_0_ − *γ/r* when the system reaches this threshold. We estimate the number of iterations to reach the threshold as

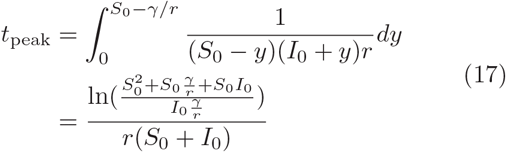

We keep the *I*_0_ explicit in this formula for clarity of units but remember *I*_0_ = 1. This is an estimate for the point in time where the random walk will lose momentum and *P*_±_ ∼ 1, leaving increase or decrease in infectives equally likely. As an aside we note that the time-to-peak is also a fat-tail distribution.[5] Ideally, *t*_peak_ should separate the two timescales discussed above. Those walks ending early will end before *t*_peak_. Those walks that reach peak infectives will end some time later, only after they dissipate nall current infectives.

Figure 2 shows how the estimate *t*_peak_ performs and compares the random walk to the standard SIR model. The vertical line in each panel indicates *t*_peak_*γ* for that choice of *S*_0_ and *I*_0_. Both *r* and *γ* are fixed. In each case, as shown in Figure 1, 1*/*ℛ_0_ is the fraction of trajectories ending before *t*_peak_*γ* time units. When ℛ_0_ = 2, 50% of the walks end before *t*_peak_*γ* yet the standard SIR model characterizes the behavior of the walks that do express an outbreak. At ℛ_0_ = 3, one third of the walks end without outbreak and at ℛ_0_ = 5 one fifth avoid outbreak. In all three of these cases the standard SIR describes the random walk behavior well and shows a slight delay in peak infectives timing as ℛ_0_ is decreased. However, when ℛ_0_ = 1.5 67% of the random walks end before *t*_peak_*γ*. This means that more than half the time there will be no outbreak at this ℛ_0_ even though ℛ_0_ > 1. This is the limit in which we would expect discrepancy between the random walk and the deterministic SIR model.

**FIG. 2:**
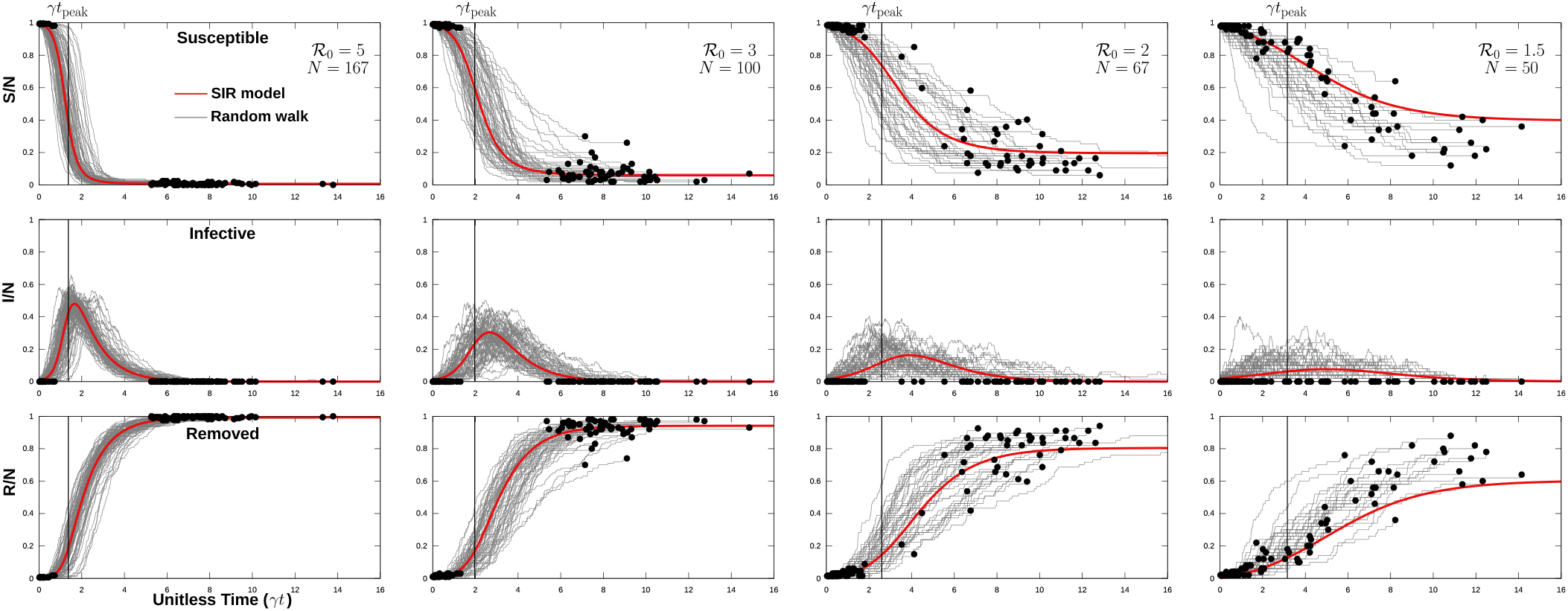
Random walk and SIR: 100 random walks are shown for fixed *r* = 6 × 10^−7^ and *γ* = 0.00002 for various population densities. Standard SIR model is shown in red for the same parameters. Circles indicate where each random walk terminated with zero infectives. Black vertical line corresponds to *t*_peak_ for each panel. Recall ℛ_0_ = *S*_0_*r/γ*.

### Application to SARS-CoV2

The above model motivates concern for fitting SARS-CoV2 data. The data at state-level is the aggregate of many counties as they encounter a common disease. Each county is in turn a composite of many communities. The above model, and the standard SIR model, are formulated on an area of uniform population density. Ideally, the model would be applied to, and fit to, real data that has been aggregated to maximize uniformity of population density. These densities may be transient in time, corresponding to gathering for an event, work, transportation, etc. When data is instead aggregated by political or taxation boundaries significant heterogeneity of population density is encountered.

To mimic this situation, we set up C = 4 individual and independent “communities” each with distinct population densities. Each community has its own initiation or lag time because each community in the SARS-CoV2 outbreak did not get seeded with its initial infection at the exact same time. Assuming that once disease is detected in an individual that individual is “removed,” then tracking the removed class of the above model would amount to tracking observed cases. Giving each community an index, *R*_*M*_ (*i*) being the *i* -th community’s cummulative case density at time *M*, ∑_*i*_ *R*_*M*_ (*i*)*A*(*i*) is the total case load in the mock “state.” Each community is assumed to present a more or less uniform density over some area *A*(*i*), which might be where people of the community work or live depending on the data and known location of transmission events.

Figure 3 displays 100 realizations of equation (6) for each of the communities, along with the “state-level” aggregate. Simulation details are given in Figure 3 caption. We solved the standard SIR model at 50 person per square mile and multiplied case density by 4 square miles to obtain the standard SIR model shown in blue in Figure 3. The standard SIR model used the exact same parameters as did each community simulation, but was performed at the state-level density. The standard SIR model will only indicate the worst possible outcome when the correct parameters are used. This owes in part to the fact that stochastic effects elevate threshold density, as discussed above and noted by others.[3, 7] Additionally, fitting to any one of the realizations in gray could produce parameter estimates that are not accurate. Concerns over fitting ℛ_0_ and ignoring the dependence on density has been raised by others[2], and here we caution against ignoring dispersion and that a distribution of outcomes is possible for a fixed parameter set.

**FIG. 3:**
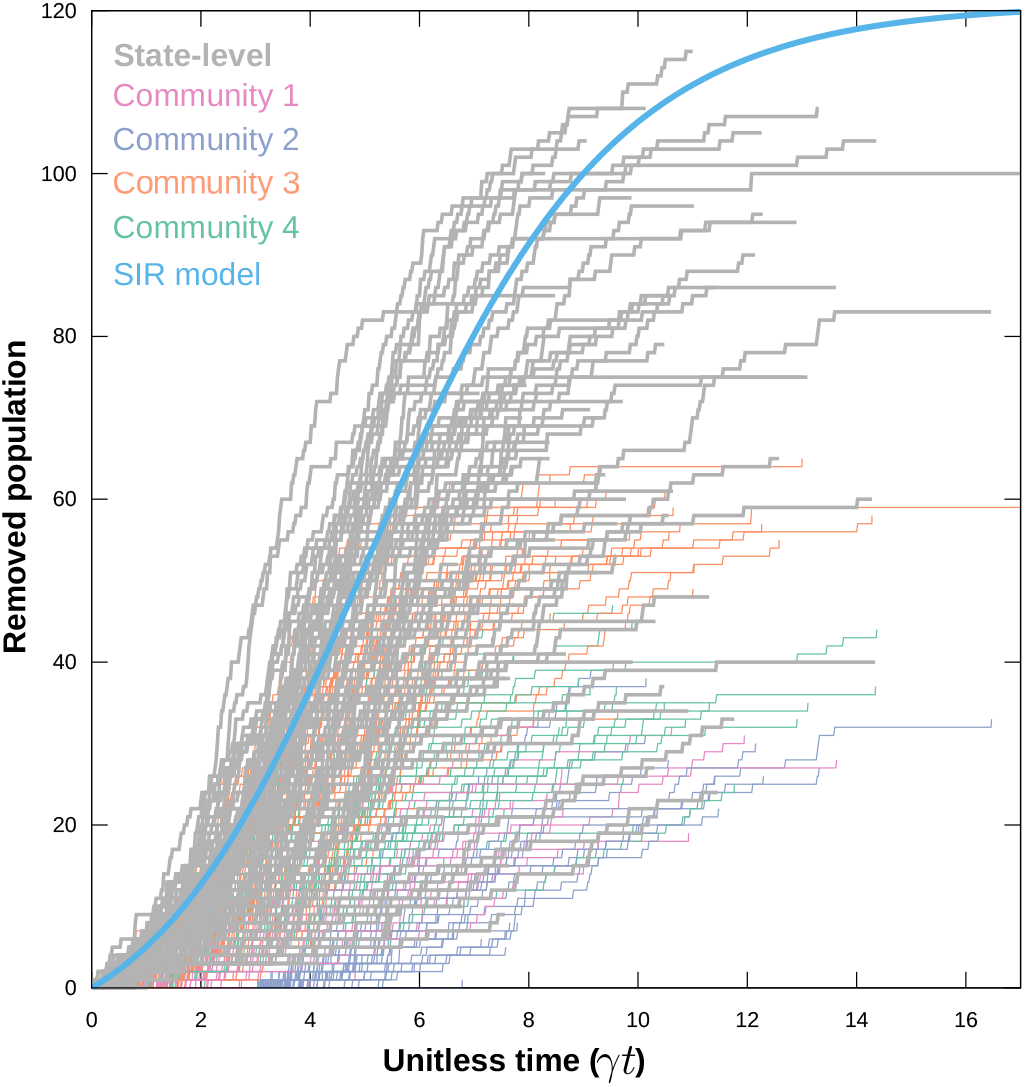
State-level vs. community-level: Community densities were: 38, 45, 67, and 50 in order. Initial seed times were: 10000, 150000, 1, and 550 iterations respectively. *r* = 6 × 10^−7^ and *γ* = 0.00002.

In Figure 1 we put forward a new way to estimate *γ/r* that does not require fitting. That procedure requires that we compute the fraction of realizations that end early as a function of population density. For the state-level aggregate shown in Figure 3, the odds of ending early are controlled by the most dense community, where there are *N* =67 people per square mile. However, any observation on the state-level outcome will be associated with the state population density of 50 persons per square mile.

For the given choice of *r* and *γ*, and Figure 2, we know ℛ_0_ = 2 for the most dense community and so the odds of ending early are 1*/*2. At the state-level, the population density is 50 persons per square mile. Given only the state-level data in Figure 3, we would measure a 50% chance of ending early. Indeed, counting the gray lines that extend above roughly 5 cases shows that only about half of the realizations displayed an outbreak of any kind. Given that half the realizations end early and a state density of 50, we obtain the estimate 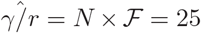. This is an incorrect estimate of *γ/r* and the error stems from using the state population density rather than the density of the most dense community. Had we used *N* = 67 for our estimate we would have found 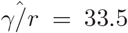, which is extremely close to the actual value of *γ/r* = 33.333.

This demonstrates that mixing heterogeneous population densities into an aggregate, such as a state or county, will produce inaccurate estimates of *γ/r*. Moreover, estimates will almost surely be an exaggeration of the true underlying values making the disease appear more aggressive than it is. This will be the case any time the state-level population density is lower than the most dense community in the state. A more accurate result could be made if the community densities are known.

Fully aware of the bias generated by aggregating data on arbitrary taxation and political boundaries, we present here an estimate of *γ/r* from SARS-CoV2 data in the United States. We collected all United States county-level data from www.usafacts.org and gathered all county population density from www.census.gov. These data were aggregated by hand and are available at github.com/cpommer/Covid19 County Data. We detected early-end outbreaks by checking for a 20 day plateau in reported cases. That is, if a county had at least 1 case and then had 20 days of zero new cases then we consider that county’s outbreak as having ended. Outbreaks may re-seed later but after 20 days of no new cases, we assume the only way to generate a new case is a re-seeding event. At the time of writing, none of the counties showing plateau by 30 days had a re-seeding event.

We tested for a 20 day plateau within the first 30, 50, 70, and 100 days since the initial case was reported. Thus, we can estimate ℱ(*t < t*^*^) for *t*^*^ = 30, 50, 70, 100 days. We suggest that plateau falling in the first 30 days are unlikely to reflect perturbation from stay-at-home orders whereas a plateau that falls in the 70 to 100 day range may well reflect perturbation from such orders. With that in mind, we expected that at the 70 to 100 day time frame the fraction of counties with “early-end” outbreaks may not follow the law expressed by equation (13). It is also observed from Figure 4(A), that beyond 40 persons per square mile there may not be enough counties at each density to allow good sampling of results.

**FIG. 4:**
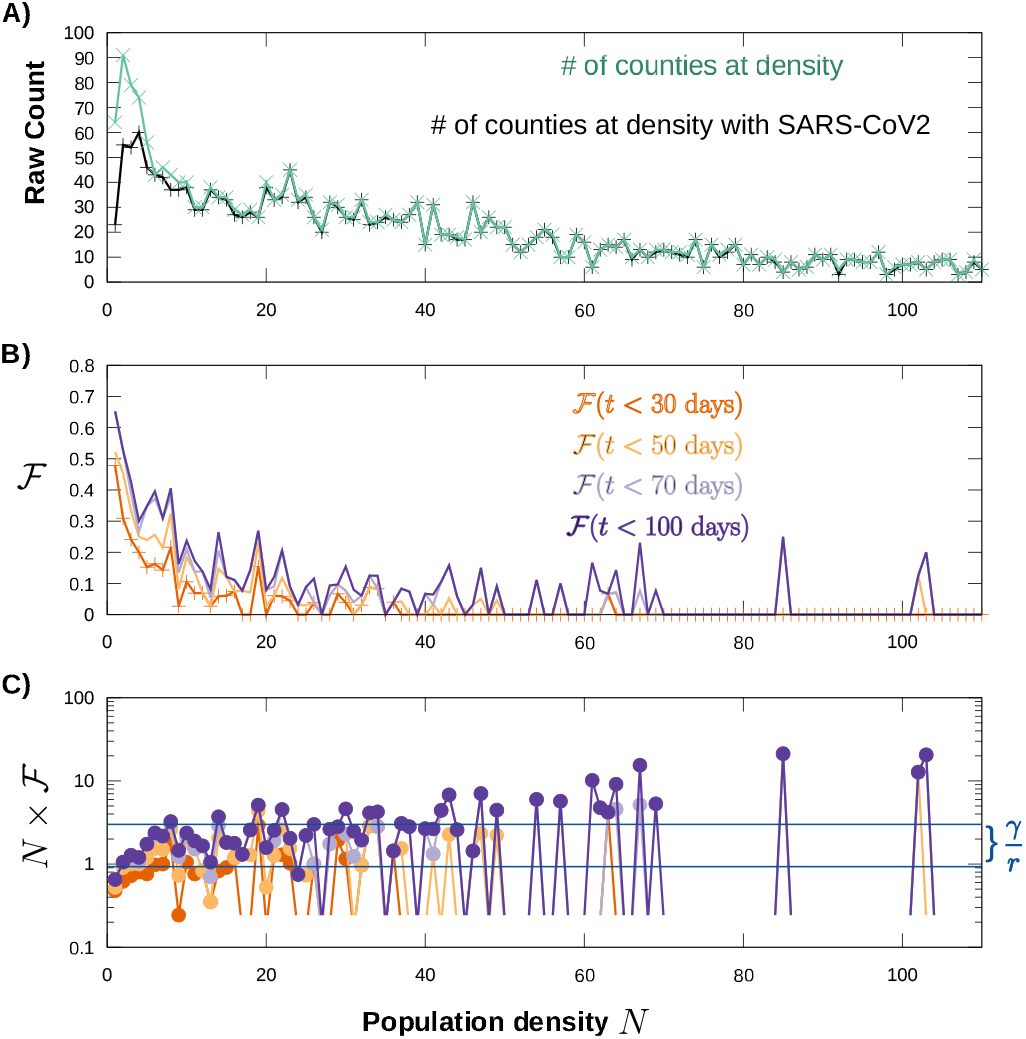
SARS-CoV2 critical density: All county data was binned on a 1 person per square mile bin-width. A) The number of U.S. counties at each population density is shown for all counties (green) and for counties that also reported SARS-CoV2 (black). B) The fraction of counties with reported SARS-CoV2 also exhibiting a 20 day plateau in cases before day 30, 50, 70, or 100. C) The critical density 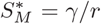 estimated as *N* × ℱ. There are not enough counties at density over 50 person per square mile for trustworthy sampling.

The county based data seems to express fairly reasonable correspondence to the model result suggesting a 1*/x* profile in ℱ (Fig. 4B). Plotting *N* × ℱ for this data we see that *γ/r* is between 1 and 3 persons per square mile (Figure 4C). This result is analogous to the model result in Figure 1B. The estimate for SARS-CoV2 is somewhat horrifying when considering the density in New York City, for example, as it suggests that if unchecked this disease will consume the susceptible population until the density of susceptibles is 1 to 3 people per square mile. However, we know that this estimate is biased.

It is trivial to argue this estimate is low by orders of magnitude. For example, New York State has a population density of around 420 people per square mile. New York County is reported to have 71886 people per square mile. Any results measured on state-level data at a density of 420 is going to be dominated by trends at the 171 times larger county density. When we measure ℱ at a county or state density, it is dominated by the highest density community (even if transient). Shifting the value of ℱ to that higher density would produce much more accurate estimates.

Unfortunately, improving our estimate of *γ/r* by applying such a shift is highly subjective. Easily, one can argue for a scaling of 100 to 1000 and obtain the range 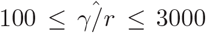, where the claim is that population mixing in grocery stores, churches, social events raises the average density during contact by up to 1000 fold. This estimate could be improved if high resolution data suitible for contact tracing were available. The large range of possible values here demonstrates how little we know about densities within counties. We noticed that San Juan county in UT, which has about 2 persons per square mile on average, has more SARS-CoV2 cases reported in the rural areas outside its most dense region. Very little can be gleaned from public data about the functional density of communities outside the areas of Blanding and Monticello, yet most of the reported cases are found outside those cities.

Our crude, subjective, estimate can be compared to a recent application of a sophisticated model for the U.S. state of Georgia.[6] Specifically, Lau and company report fitted estimates of ℛ_0_ in Cobb, Dekalb, Fulton, Gwinett, and Dougherty counties. Since they also estimate a strong degree of dispersion and ‘super-spreading’ in those counties we understand these ℛ_0_ values as a sort of average taking only a representative value. Since *γ/r* = *N* × ℱ = *N/*ℛ_0_, we can use county density and their estimates of ℛ_0_ to validate our crude estimate of 100 ≤ *γ/r* ≤ 3000. In order, census data gives the following population densities for those counties: 2203, 2777, 1940, 2096, 288 persons per square mile. We note that the city of Albany in Dougherty county has a density of 1354 persons per square mile. These densities combined with the fits of ℛ_0_ reported by others[6] produce the following critical density estimates in order: 759, 925, 808, 1103, 58 persons per square mile. Note that in Dougherty county we obtain the estimate 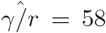 at the county density of 288 and 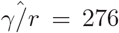 using the density of Albany city, again showing that population dispersion is important. The cases in Dougherty county have largely been rationalized as resultant to a “superspreading” event, where a single infective interacts with suceptibles in a situation where population density is extremely high. With an average of 11 feet between people, population density is 76923 persons per square mile. With an average of 3 feet between people the density jumps to 10^6^ persons per square mile. Super-spreading events likely occur at this later density where people are mingling and interacting. These transient, high-density situations (like special events) make it impossible to accurately evaluate *γ/r* with only county and state-level population densities.

Our approach does not use fitting, but rather categorizes data by population density and measures the number of communities (or counties in this case) that have early-end outbreaks. Limited by the accuracy of county-level population density, our estimate falls in rough agreement with the fitted ℛ_0_ values (where in theory ℛ_0_ = *S*_0_*r/γ*) obtained by others[6] fully acknowledging the subjective nature of correcting county densities. With better resolution of transient organization of population density it may be that *γ/r* is actually in the tens of thousands, consistent with the current view that super-spreading plays a key role in the SARS-CoV2 outbreak.[6] One can imagine that if SARS-CoV2 case status was linked to anonymized cell phone data, highly accurate acounting of transient population density and super-spreading events would emerge. Calculating ℱ then arises as an accurate way to estimate key parameters directly from data without fitting to models. Currently, we are limited to public data which only reports cases at the resolution of county and we have no way to account for the dispersion of population within county boundaries.

Equation (17) cannot exactly be applied to the current pandemic because we do not have separate accounts of *r* and *γ*. Moreover, the time-to-peak is more sensitive to perturbation by stay-at-home order. However, there is a clear dependence on density in the time-to-peak, with higher densities generally reaching peak sooner than lower densities. This provides some basis for understanding the large distribution of peak-times being observed across the United States, even though the observed times are sure to be further perturbed by stay-at-home orders and changing opinions as to whether or not those orders should be followed.

## CONCLUSION

We have put forward a random walk that is on average consistent with the standard SIR model. The key interpretations of the standard model were introduced for the random walk. This led directly to a new way to estimate the critical population of susceptibles defining the threshold of outbreak without requiring model fitting. We used this technique to estimate the critical threshold for the SARS-CoV2 outbreak in the United States based on county-level data, and made light of the associated caveats. We also rationalize the disparities in time-line of the outbreak through equation (17), which gives an estimate for the time of peak infectives.

## CODE AND DATA AVAILABILITY

The code, gnuplot scripts, and data presented above can be found at GitHub github.com/BradleyDickson/ideal gas and sir

## Data Availability

All code and data are available on GitHub, links are in the manuscript and below.

https://github.com/BradleyDickson/ideal_gas_and_sir

## ACKNOWLEDGEMENTS

This work was supported by the Van Andel Research Institute’s Staff Scientist mechanism. We thank Chris Pommer for providing data and entertaining several discussions. Robert M. Vaughan and Evan M. Cornett are thanked for editorial help. We also thank Arthur F. Voter for his comments and insightful discussions.

## Notes

### Competing Interest Statement

The authors have declared no competing interest.

### Author Declarations

does not apply

## References

[1] William Ogilvy Kermack and Anderson G McKendrick. A contribution to the mathematical theory of epidemics. Proceedings of the royal society of london. Series A, Containing papers of a mathematical and physical character, 115(772):700–721, 1927.

[2] Claire Donnat and Susan Holmes. Modeling the heterogeneity in covid-19’s reproductive number and its impact on predictive scenarios. arXiv preprint arXiv:2004.05272, 2020.

[3] Laurent Hébert-Dufresne, Benjamin M Althouse, Samuel V Scarpino, and Antoine Allard. Beyond r0: Heterogeneity in secondary infections and probabilistic epidemic forecasting. medRxiv, 2020.

[4] Mark Buchanan. The limits of a model. Nature Physics, 16, 2020.

[5] Pasquale Cirillo and Nassim Nicholas Taleb. Tail risk of contagious diseases. Nature Physics, pages 1–8, 2020.

[6] Max SY Lau, Bryan Grenfell, Kristin Nelson, and Ben Lopman. Characterizing super-spreading events and agespecific infectivity of covid-19 transmission in georgia, usa. medRxiv, 2020.

[7] Roy M Anderson and Robert M May. Infectious diseases of humans: dynamics and control. Oxford university press, 1992.

[8] Gary F Hatke, Monica Montanari, Swaroop Appadwedula, Michael Wentz, John Meklenburg, Louise Ivers, Jennifer Watson, and Paul Fiore. Using bluetooth low energy (ble) signal strength estimation to facilitate contact tracing for covid-19. arXiv preprint arXiv:2006.15711, 2020.

[9] Hao Hu, Karima Nigmatulina, and Philip Eckhoff. The scaling of contact rates with population density for the infectious disease models. Mathematical biosciences, 244(2):125–134, 2013.

[10] Daryl J Daley and Joe Gani. Epidemic modelling: an introduction, volume 15. Cambridge University Press, 2001.

[11] Paul L Delamater, Erica J Street, Timothy F Leslie, Y Tony Yang, and Kathryn H Jacobsen. Complexity of the basic reproduction number (r0). Emerging infectious diseases, 25(1):1, 2019.

[12] Marc Lipsitch, Ted Cohen, Ben Cooper, James M Robins, Stefan Ma, Lyn James, Gowri Gopalakrishna, Suok Kai Chew, Chorh Chuan Tan, Matthew H Samore, et al. Transmission dynamics and control of severe acute respiratory syndrome. Science, 300(5627):1966– 1970, 2003.

[13] Ryogo Kubo and Donald A McQuarrie. Statistical mechanics. PhT, 18(10):74, 1965.

[14] Roberto Mauri. Non-Equilibrium Thermodynamics in Multiphase Flows. Springer Science & Business Media, 2012.

